# Deprescribing Pro Re Nata Antihypertensives in the Hospital An Implementation Science Pilot Study Based on the COM-B Framework

**DOI:** 10.1101/2025.08.18.25333937

**Authors:** Anthony Fangman, Srilatha Gannavaram, Mark Steinbeck, Rebecca Adler, Abdul Wali Khan, Zhuxuan Fu

**Affiliations:** Hospital Medicine Division, Saint Luke’s Physicians Group; Pharmacy Division, Saint Luke’s Health System; Department of Internal Medicine, University of Missouri Kansas City School of Medicine; Biostatistics, Saint Luke’s Research Administration

## Abstract

Elevated blood pressure is a common occurrence in acute care settings irrespective of the reason for hospitalization. A large body of available evidence suggests possible harm from intensive hypertension management when there is no evidence of end-organ damage, otherwise known as asymptomatic hypertension (HTN). Despite this, Pro Re Nata (PRN) antihypertensives are commonly prescribed to hospitalized adults and utilized to treat asymptomatic hypertension. This practice contributes to patient harm and is considered low value care. So far, there is limited evidence to guide interventions that can successfully and consistently deprescribe PRN antihypertensive use in acute care settings. To address this gap, we conducted a pilot study in which we developed an intervention bundle using implementation science concepts based on the COM-B framework. Our results showed a significant decrease in the use of PRN antihypertensive prescriptions for hospitalized adults by hospitalist providers, especially following the delivery of high-fidelity interventions, This pilot was conducted in a single health care system comprising multiple hospitals. Due to their low cost and easy adaptability, we believe that the methods described here can be reproduced widely to minimize patient harm and allow ongoing evaluation of practices to address elevated blood pressure in acute settings. Further, this study demonstrates the feasibility of integrating implementation science principles into responsible management of hypertension among hospitalized adults based on available evidence.

## INTRODUCTION

Hypertension is a chronic disease condition among adults and a well-recognized risk factor for various adverse health outcomes including coronary artery disease, stroke, renal disease, and increased mortality (24,25). In primary care, appropriate recognition of this condition and evidence-based treatment have proven benefits in improving health outcomes (24,25). On the other hand, treatment of hypertension in the inpatient setting is much more complicated due to the lack of established guidelines (23). Multiple studies in fact demonstrated harm when HTN is aggressively treated in the hospital except when a hypertensive crisis with end-organ damage is present (2-6,8,10,12-14,18). The reasons for such disparity in outcomes between ambulatory and acute settings are likely to be multifold. Possible factors include spurious blood pressure elevations, inappropriate timing and/or technique of blood pressure measurements, IV fluids, fluid overload, glucocorticoids or other treatments that could transiently raise blood pressure, missed home medications, patient anxiety, inadequate pain control, adrenergic response associated with acute illnesses and others that are yet to be identified (15-17). Therefore, it is very important to distinguish between asymptomatically elevated blood pressures and hypertensive emergencies among hospitalized adults. The former requires an assessment of causes followed by a tailored treatment plan whereas the latter requires initiation of immediate treatment with established protocols based on the type of end-organ damage (5). Scientific statement from American Heart Association states that asymptomatically elevated blood pressure in the inpatient setting should not be treated routinely (13). Most importantly, PRN antihypertensive use has been linked to patient harm and adverse effects including hypotension, acute kidney injury, strokes, increased length of stay and mortality among hospitalized adults (2-6,8,10,12-15,18).

The prevalence of elevated blood pressure in adult medical inpatients has been estimated to be as high as 72% (1). However, only 0.6% of patients that present to acute care facilities without evidence of end-organ damage are estimated to have a hypertensive emergency (5). Despite this, the use of PRN antihypertensives remains a widely prevalent practice affecting an estimated 21-35% of all hospitalized adults (13,28). Routine prescriptions for PRN antihypertensives in the hospital are problematic for three reasons. First and foremost, blood pressure is treated without giving providers an opportunity to assess whether end-organ damage is present, potentially leading to inappropriate treatment of hypertensive emergencies. Secondly, asymptomatically elevated blood pressures are aggressively treated, increasing patient risk. Lastly, this practice results in an increased utilization of low value care and high costs associated with unnecessary treatment.

A prior Quality Improvement initiative by hospitalist providers successfully decreased the use of IV Hydralazine and Labetalol among patients with elevated blood pressures without increasing patient harm (11). In our study, we sought to expand these interventions, broaden the number of PRN antihypertensives targeted, and aimed to avoid extensive patient-level data acquisition in order to increase adaptability among various acute care facilities. To this end, we used implementation science concepts based on the COM-B framework that included assessment of barriers followed by the development of an intervention bundle. Our primary outcome measure was a reduction in the percentage of patients with prescriptions for PRN antihypertensives. Additional outcome measures were a reduction in the total number of PRN antihypertensive orders by hospitalist providers, a sustained change in provider prescribing patterns and responsiveness of outcomes to the fidelity of interventions. We also monitored the use of medication orders to treat hypertensive emergencies, and deaths among patients treated for hypertensive emergencies to measure the potential for unintended harms associated with a reduction in antihypertensive medications.

## METHODS

As we considered interventions that can be successfully implemented and sustained, we sought to identify those that involved no cost or low cost to the hospitalist division and the health system, use Electronic Health Record (EHR) data to which our hospitalist division has ready access and avoid collection of extensive patient-level data.

The study was conducted in all Saint Luke’s Health System (Barnes Jewish West Region) hospitals where hospitalists manage patients. Stakeholders for the study were all hospitalist providers within the health system. We received a Quality Improvement determination from our Internal Review Board.

We used the COM-B framework, and the Behavior Change Wheel (27) to perform a barrier assessment. Per semiformal communications with hospitalist providers and a priori knowledge, the following barriers were identified for de-implementation of PRN antihypertensives in the acute care setting:

1. Provider lack of knowledge regarding potential harms of aggressive treatment of asymptomatic HTN.
2. Provider reluctance to change practice due to perceived harm to patients caused by elevated blood pressure.
3. Nursing expectations for providers to place PRN orders when blood pressure is elevated.
4. Inadequate time for providers to consider alternative explanations and develop a tailored treatment plan.
5. Provider desire to maintain autonomy over patient-care decisions regarding blood pressure management.
6. Provider aversion to frequent Best Practice Advisory (BPA) alerts built into Electronic Health Record (EHR).

Following barrier assessment, we identified three intervention areas-Education, Environmental restructuring and Persuasion to develop the following intervention bundle.

### Intervention Bundle 1 – Education

Consisted of (i) group education and (ii) tailored education to providers with PRN antihypertensive orders higher than group average.

### Intervention Bundle 2 - Environmental restructuring

Consisted of (i) updating hospitalist admission order set with a higher blood pressure threshold for nursing staff to alert providers if patients did not have new symptoms and (ii) engaging nursing leadership to influence nursing expectations for PRN antihypertensives. Formal nursing education was not provided.

### Intervention Bundle 3 – Persuasion

Consisted of two audit and feedback interventions and reminders. We collected monthly data on the number of PRN antihypertensive orders generated by individual hospitalist providers. This data was used to provide (i) feedback by site (number of orders generated at each hospital site were shared with all providers at that site), (ii) targeted feedback by provider (providers prescribing higher than group average were sent tailored education). Providers identified for targeted feedback were made aware that their PRN antihypertensive use was higher than the group average. They were provided with links to relevant scientific literature. In addition, they were informed that chart reviews were not done, and that the appropriateness of prescriptions has been deferred to individual providers caring for patients. Lastly, reminders were provided to all hospitalist providers regarding the reasons we were undertaking these interventions.

We utilized a quasi-experimental interrupted time series design with our research plan being carried out over six periods including baseline as described below. Each period consisted of three consecutive months.

i. June 2023-August 2023 **(Baseline) - No intervention**. Retrospective data collection.
ii. Sept 2023-November 2023 **(Period 1) - Intervention: General provider education**. All providers in the hospitalist division of our health system were provided education about potential harms of using PRN antihypertensives.
iii. January 2024-March 2024 **(Period 2) - Intervention: Environmental restructuring**. See above for description.
iv. April 2024-June 2024 **(Period 3)-Intervention: Audit and feedback by site**. See above for description.
v. July 2024-September 2024 **(Period 4) - Intervention: Audit and targeted feedback (tailored education)**. See above for description.
vi. October 2024-December 2024 **(Period 5) - No intervention**. Ongoing monthly data collection.

Fidelity to various interventions varied from medium to high, as indicated in Table 1. Listed interventions were carried out in addition to modeling targeted behavior by medical directors and selected senior partners in the group which was ongoing prior to and throughout the study period.

**Table 1:** Interventions by period and fidelity to each intervention.

| **Baseline** | **Period 1** | **Period 2** | **Period 3** | **Period 4** | **Period 5** |
| --- | --- | --- | --- | --- | --- |
| **Jun-Aug 2023** | Sep - Nov<br>2023 | Jan - Mar<br>2024 | Apr - Jun<br>2024 | Jul - Sep<br>2024 | Oct - Dec<br>2024 |
| **No intervention** | **Education** | **Environmental restructuring** | **Audit & feedback by site** | **Audit & targeted feedback** | **No intervention** |
| **Retrospective data collection** | Group meetings, emails<br><u>(Medium Fidelity)</u> | Discussions with nursing leadership<br><u>(Medium Fidelity)</u><br>Modified order set with higher BP threshold<br><u>(High Fidelity)</u> | Monthly data collection. Distribute PRN use by site.<br><u>(High Fidelity)</u><br>Group reminders to avoid aggressive HTN treatment if not HTN emergency<br><u>(Medium Fidelity)</u> | Monthly data collection. Identify high users (>5 PRN orders/month)<br>Individual message to high users with targeted education<br><u>(High Fidelity to all)</u> | Ongoing monthly data collection |

All data points were extracted using the Slicer Dicer function in our health system’s EHR, Epic. Data was then exported to Microsoft Excel with security features and analyzed.

## OUTCOMES

We collected the following data to monitor outcomes. All data were collected at the end of each period except #1, which was collected monthly.

1. Number of PRN antihypertensive orders by site and by provider for the following medications
2. IV Hydralazine, PO Hydralazine, IV Labetalol, PO Clonidine, IV Enalaprilat.
3. Number of orders for the following infusions to treat HTN emergency (not limited to orders by hospitalist providers).
4. Esmolol, Nicardipine, Nitroglycerin, Phentolamine and Sodium Nitroprusside.
5. Number of deaths among patients that received an infusion for HTN emergency.
6. Number of unique patient encounters by hospitalist providers

The above data was analyzed to evaluate changes in the percentage of patients with PRN antihypertensive orders, overall numbers and distribution of PRN antihypertensives, provider prescribing patterns, effects on the number of infusions ordered for hypertensive emergencies and deaths among patients that received at least one infusion for hypertensive emergency.

### Percentage of PRN antihypertensives per patient encounter

The number of unique hospitalist encounters for each period during the study varied from 8883 to 9909. The percentage of encounters with PRN antihypertensive medication orders was calculated using this data combined with PRN antihypertensive orders data. We found that the percentage of encounters with PRN antihypertensive orders decreased from 14.7% at baseline to 2.7% in period 5, with the most significant reduction occurring in the use of IV Hydralazine from 10.5% of patient encounters to just 0.74%. These results reflect a substantial reduction in the percentage of patients treated by hospitalists that had prescriptions for PRN antihypertensives.

### Individual Provider PRN antihypertensive prescribing patterns

Prescription patterns of individual providers were analyzed, omitting those providers that were not present for the entirety of the study period. Each of these ninety-two providers were placed in one of five groups based on the number of PRN antihypertensive orders during the preceding period. The groups were (i) Extremely high: greater than 50 orders, (ii) High: 21-50 orders, (iii) Moderate: 11-20 orders, (iv) Low: 6-10 orders and (v) Minimal: Less than 5 orders.

At baseline, there were two providers in the extremely high group, fifteen in the high group, thirty-eight in the moderate group, seventeen in the low group and twenty in the minimal group. At the end of period five, none of the providers were in extremely high or high groups. One provider was in the moderate use group; seven providers were in the low use group and the remaining eighty-four providers were in the minimal use group (Table 2). We observed a reduction in the number of providers in the high, moderate and low use groups following high-fidelity interventions (order set modification, audit and feedback) in the period preceding data collection. The number of providers in the minimal use category increased following the delivery of high-fidelity interventions in the period preceding data collection (Figure 1).

**Table 2:** Provider groups by period.

| Periods | Orders |  |  |  |  |  |
| --- | --- | --- | --- | --- | --- | --- |
|  | Extremely High<br>(> 50) | High<br>(21-50) | Moderate<br>(11-20) | Low<br>(6-10) | Minimal<br>(≤ 5) | Total |
| Baseline | 2 | 15 | 38 | 17 | 20 | 92 |
| Period 1 | 2 | 12 | 31 | 24 | 23 | 92 |
| Period 2 | 1 | 8 | 30 | 20 | 33 | 92 |
| Period 3 | 1 | 4 | 10 | 28 | 49 | 92 |
| Period 4 | 0 | 1 | 9 | 7 | 75 | 92 |
| Period 5 | 0 | 0 | 1 | 7 | 84 | 92 |

**Figure 1:**
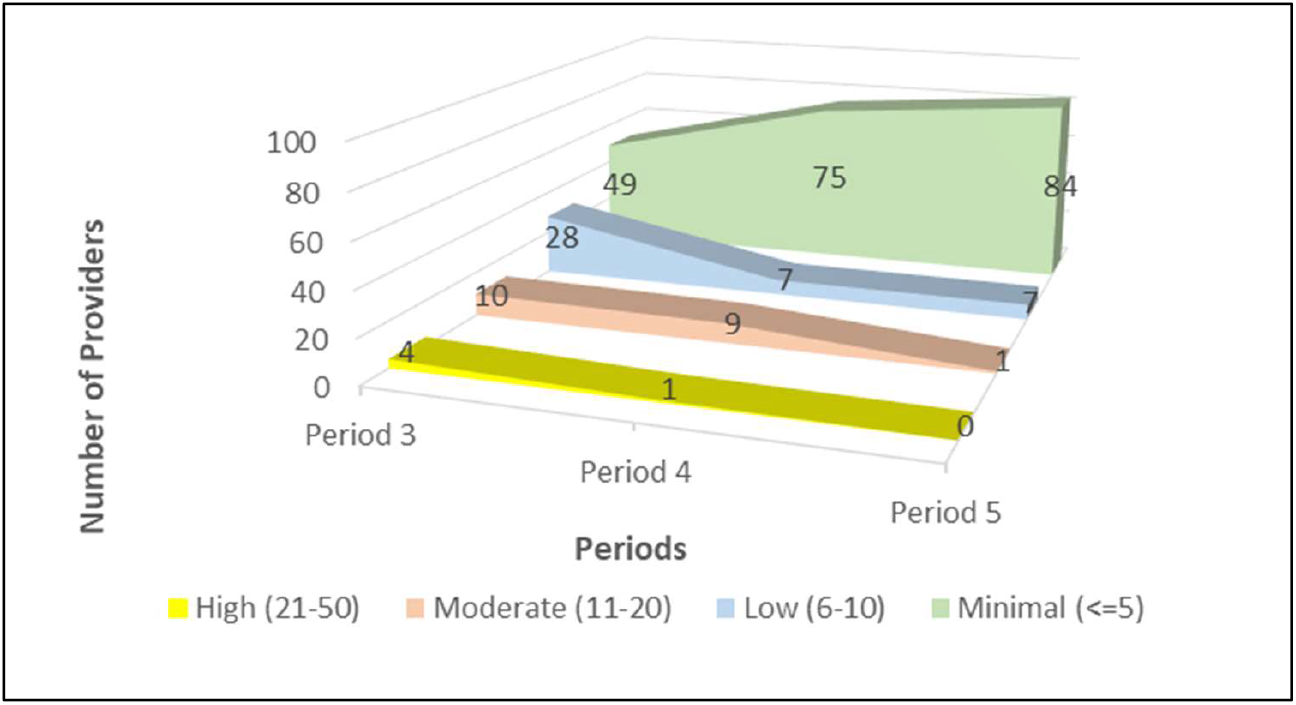
Provider grouping following high fidelity interventions

### Effects on the number of total PRN antihypertensives

During the baseline period, 1329 PRN antihypertensive orders were placed by hospitalist providers. For the subsequent two periods of 3 months each, a slight reduction of overall PRN antihypertensives and IV hydralazine was noted. Substantial reductions were noted in both overall PRN antihypertensive use and IV hydralazine use in periods 3 through 5 (Table 3).

**Table 3:** PRN antihypertensive order trends.

| PRN orders | Baseline | Period 1 | Period 2 | Period 3 | Period 4 | Period 5 |
| --- | --- | --- | --- | --- | --- | --- |
| **Grand total** | **1329** | **1200** | **1059** | **708** | **444** | **275** |
| **IV hydralazine** | 951 | 780 | 623 | 327 | 101 | 74 |
| **PO hydralazine** | 66 | 83 | 74 | 59 | 32 | 26 |
| **Labetalol** | 238 | 243 | 270 | 238 | 224 | 131 |
| **Clonidine** | 67 | 85 | 83 | 70 | 76 | 33 |
| **Enalaprilat** | 7 | 8 | 9 | 14 | 11 | 11 |

### Effects on the distribution of PRN antihypertensive orders

IV Hydralazine was the predominant PRN antihypertensive in the baseline and subsequent two periods, accounting for 71.5%, 65% and 58.8% of overall PRN antihypertensives, respectively. In period 3, IV hydralazine decreased to 46.1% and further decreased in periods 4 and 5 to 22.7% and 26.9% respectively. IV labetalol percentage rose from 17.9% at baseline to 47.6% of the overall PRN antihypertensives in period 5, becoming the most common PRN IV antihypertensive prescribed by hospitalist providers at the end of period 5 (Figure 2). A review of the total number of these prescriptions demonstrates that while the percentage of labetalol use increased, the overall number of orders decreased from 238 in baseline period to 131 in period 5.

**Figure 2:**
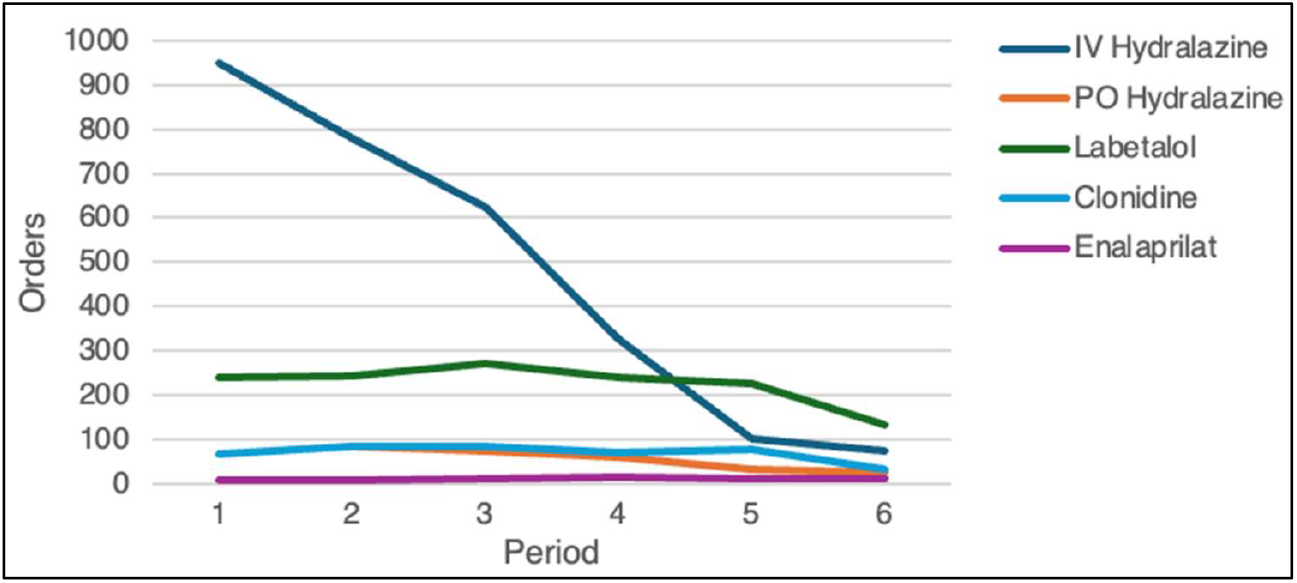
Number and types of PRN antihypertensive orders by period

### Treatment of Hypertensive Emergencies

We collected data on the number of infusions and medications used for hypertensive emergencies throughout our health system for the study period. These included Esmolol, Nicardipine, Nitroglycerin, Phentolamine, and Sodium Nitroprusside. We excluded procedural use of these medications and included their use in any department or any provider, not limiting use by hospitalist providers. Our results showed that despite a conservative approach to hypertension management, there was a reduction in the overall use of medications administered for hypertensive emergency treatment (Table 4). A review of the number of deaths that occurred among patients that received one or more of these medications for hypertensive emergency management did not show meaningful change.

**Table 4:** Number of Infusions to treat HTN emergencies.

| **Infusions** | **Period 1** | **Period 2** | **Period 3** | **Period 4** | **Period 5** | **Period 6** |
| --- | --- | --- | --- | --- | --- | --- |
| Esmolol | 23 | 15 | 16 | 20 | 12 | 32 |
| Nicardipine | 306 | 359 | 324 | 311 | 314 | 351 |
| Nitroglycerine | 159 | 111 | 77 | 64 | 68 | 83 |
| Phentolamine | 5 | 4 | 11 | 3 | 7 | 10 |
| Sodium Nitroprusside | 67 | 56 | 71 | 71 | 71 | 53 |
| Grand Total | 560 | 545 | 499 | 469 | 472 | 529 |
| # HTN emergency deaths | 25 | 26 | 24 | 17 | 25 | 28 |

## DISCUSSION

Elevated blood pressure is a common occurrence among hospitalized patients. Many of these patients do not have evidence of end-organ damage yet frequently receive treatment with PRN antihypertensives that are administered based on blood pressure values alone. This practice can increase patient harm as a large body of observational data has shown that the use of PRN antihypertensives can increase the risk of adverse patient outcomes. Use of PRN antihypertensives without appropriate evaluation to assess the presence of end-organ damage may lead to undiagnosed incidences of hypertensive emergencies. In addition, a majority of PRN antihypertensives are IV medications and are associated with increased expenditures. For these reasons, it is vital to develop strategies that curb the routine prescription of PRN antihypertensives for adult inpatients. Current American Heart Association guidelines encourage clinicians to consider possible causes for blood pressure elevation and develop an individualized approach for management.

In this pilot study, we utilized an implementation science framework with an aim to deprescribe PRN antihypertensives among hospitalized adult patients. Using a COM-B framework, we performed a barrier analysis followed by the development of an intervention bundle broadly consisting of education, environmental restructuring, and persuasion. Our results demonstrate a significant decrease in the percentage of patients receiving orders for PRN antihypertensives. In addition, we noted a reduction in the overall number of PRN antihypertensive orders, especially IV hydralazine. Despite a reduction in PRN antihypertensive orders, we noted a slight reduction in the number of infusions for hypertensive emergency management system-wide and no change in mortality among patients treated for hypertensive emergency.

A significant advantage of our study is that our interventions did not involve any cost to the hospital medicine division or our health system and did not rely on extensive patient-level data collection. Therefore, we believe that these interventions can be easily adapted to various acute care facilities. In addition, our approach would allow possible refinement of interventions as needed based on outcome measures or available evidence regarding blood pressure management in the hospital.

We believe that our study adds to existing knowledge regarding the management of hypertension in acute care settings in several important ways. First and foremost, this is the first study to successfully incorporate implementation science concepts to address PRN antihypertensive use in hospitals. Use of the COM-B framework to influence provider prescribing practices in hospital medicine is also novel. Implementation science has the potential to bridge the gap between rapidly evolving evidence and increasingly complex clinical practice. This study demonstrates the feasibility and ability of carefully selected implementation science concepts to attain meaningful outcomes in hypertension research. Second, the described interventions can be easily sustained and/or replicated owing to their favorable costs and easy reproducibility. This would allow comprehensive data collection and measurement of outcomes that will deepen our knowledge about hypertension management in the inpatient setting. Lastly, we believe that our study has demonstrated an effective and responsible approach to the use of widely available EHR data extraction tools that can be leveraged to design, monitor, and adapt interventions for appropriate hypertension management for hospitalized adults.

Caution must be exercised in interpreting these results. Lack of randomized trial evidence on how to manage hypertension in the inpatient setting makes it challenging to offer specific guidance to providers. Despite careful messaging, the proposed interventions may create confusion among clinicians regarding the use of antihypertensives in appropriate clinical settings. In our study, we took care to reinforce the message that PRN antihypertensives should be minimized only in the treatment of asymptomatic HTN. EHR-generated reports may contain errors despite careful extraction and need to be interpreted cautiously. Lastly, it is unknown if provider behaviors will continue to respond to these interventions, have reached a plateau or revert to pre-intervention patterns. We plan to continue ongoing audits to analyze the longevity of these results and consider additional implementations such as a peer review of providers with ongoing high PRN use, sharing unblinded provider data with all hospitalist providers or refining provider education.

Future studies will be needed to fully evaluate the reproducibility of our methods and results along with a thorough evaluation of potential harms caused by untreated asymptomatic hypertension in the hospital setting. Ultimately, randomized controlled trials will be needed to establish the best treatment strategies for elevated blood pressure in the inpatient setting.

## Data Availability

Data used in this study will be made available to reviewers after removal of any individually identifiable information.

